# Experiences of families using an early example of neighbourhood multidisciplinary care for children and young people: A qualitative exploration using a theoretical framework of acceptability

**DOI:** 10.64898/2026.05.14.26353240

**Authors:** I Litchfield, F Dutton, L Harper, S Kaur, C Luxmore, L Rahman, C Wolhuter, C Bird

**Affiliations:** Department of Applied Health Research, University of Birmingham, UK; Birmingham Health Partners, Birmingham, UK; Small Heath Medical Practice, Birmingham, UK; GreenSquareAccord, UK; Birmingham Women’s and Children’s NHS Foundation Trust, Birmingham, UK; Nuffield Department of Primary Care Health Sciences, University of Oxford, UK

**Keywords:** Child health, Qualitative research, Health policy, Health services research

## Abstract

**Background:** In the United Kingdom, the National Health Service is attempting to address the ongoing challenges to heath equity in underserved children and young people (CYP) by creating Neighbourhood Multi-Disciplinary Teams (NMDTs) that combine health services, social care providers, local authorities, voluntary, community and faith and social enterprise is needed. Despite this significant shift in the delivery of care, there is a lack of suitably robust evidence of family experience to inform their development. This work contributes to this need using the experience and perspectives of families using an early example of an NMDT for CYP; the “Sparkbrook Children’s Zone” in Birmingham (UK).

**Methods:** The study used data collected from two focus groups conducted with parents whose children had been treated by the Sparkbrook Children’s Zone. The data were analysed using a directed content analysis to populate Sekhon’s Theoretical Framework of Acceptability.

**Results:** In summary (by Framework domain) we found that that individuals became aware of the SCZ through a range of sources, understanding that it was multidisciplinary if sometimes unsure of precisely the organisations involved (*Intervention coherence*); Parents described the benefits to access of a locally situated collocated service (*Burden*) the personalised relationship with providers (*Cultural sensitivity*), extended consultation time, and support for the family’s complex clinical and social needs (*Perceived effectiveness*).

**Conclusions:** Parents appeared to prefer the SCZ over usual primary care but more work is needed with larger sample sizes to ensure that the structure of NMDTs are understood and optimised.

**Key messages:**

- In the United Kingdom the rapid implementation of multidisciplinary primary care for children and young people has begun despite a lack of robust evidence
- This study provides valuable insight into the experiences and preferences of families using an early example of a neighbourhood multidisciplinary care team in a socio-economically challenged ward in Birmingham.
- The collocated combination of clinical and social support was welcomed as was the personalised nature of the clinical care and the value of the practical support and advocacy to address families’ complex social needs.

**Patient or public contribution:** A patient and public involvement panel was created when the Sparkbrook Children’s Zone was first developed, and they continued to offer advice on the evaluation of the service. They were involved in the decision to collect data via focus groups and supported the development of the focus group topic guides, and the language used in the recruitment materials. The early results were fed back to the PPI panel and they are supporting the dissemination of the evaluation findings through their affiliation with local community and faith groups.

## Introduction

Policymakers and commissioners in the United Kingdom are recognising that a more holistic approach is needed to address the ongoing challenges to heath equity in underserved CYP [1, 2]. The most recent National Health Service (NHS) Long Term Plan has attempted to meet this need through the introduction of Neighbourhood Multi-Disciplinary Teams (NMDTs) [3]. These contain primary-care led teams integrated with wider services including education, social support (including social care), and the Voluntary Community, Faith and Social Enterprise (VCFSE) sector [4, 5]. They are expected to better meet the needs of CYP in underserved populations, reducing referrals to secondary care, and increasing opportunities for early intervention and preventive health [3]. Despite this significant shift in the delivery of care there is a lack of suitably robust evidence to inform these complex service-level interventions [6], this includes an understanding of the concerns, preferences, and experiences of target populations [7].

One early example of an NMDT that offers the opportunity to understand the experiences of CYP and their families is the Sparkbrook Children’s Zone. The service collocates general practitioners, family support workers, mental health outreach, and paediatricians in a low-income area of Birmingham (UK) to deliver placed-based health and social care to underserved CYP (see Figure 1 for the service blueprint) [8]. To complement the previous studies that have described the health economic impact of the service [9], staff experience [10, 11], and service utilisation [12], the work presented here presents the findings of a qualitative exploration of the perspectives and preferences of families that have used the service. The data were analysed and presented through an *a priori* framework of acceptability [13], to provide structured insight into their awareness and understanding of the service, its accessibility, cultural alignment and perceived effectiveness.

**Figure 1.**
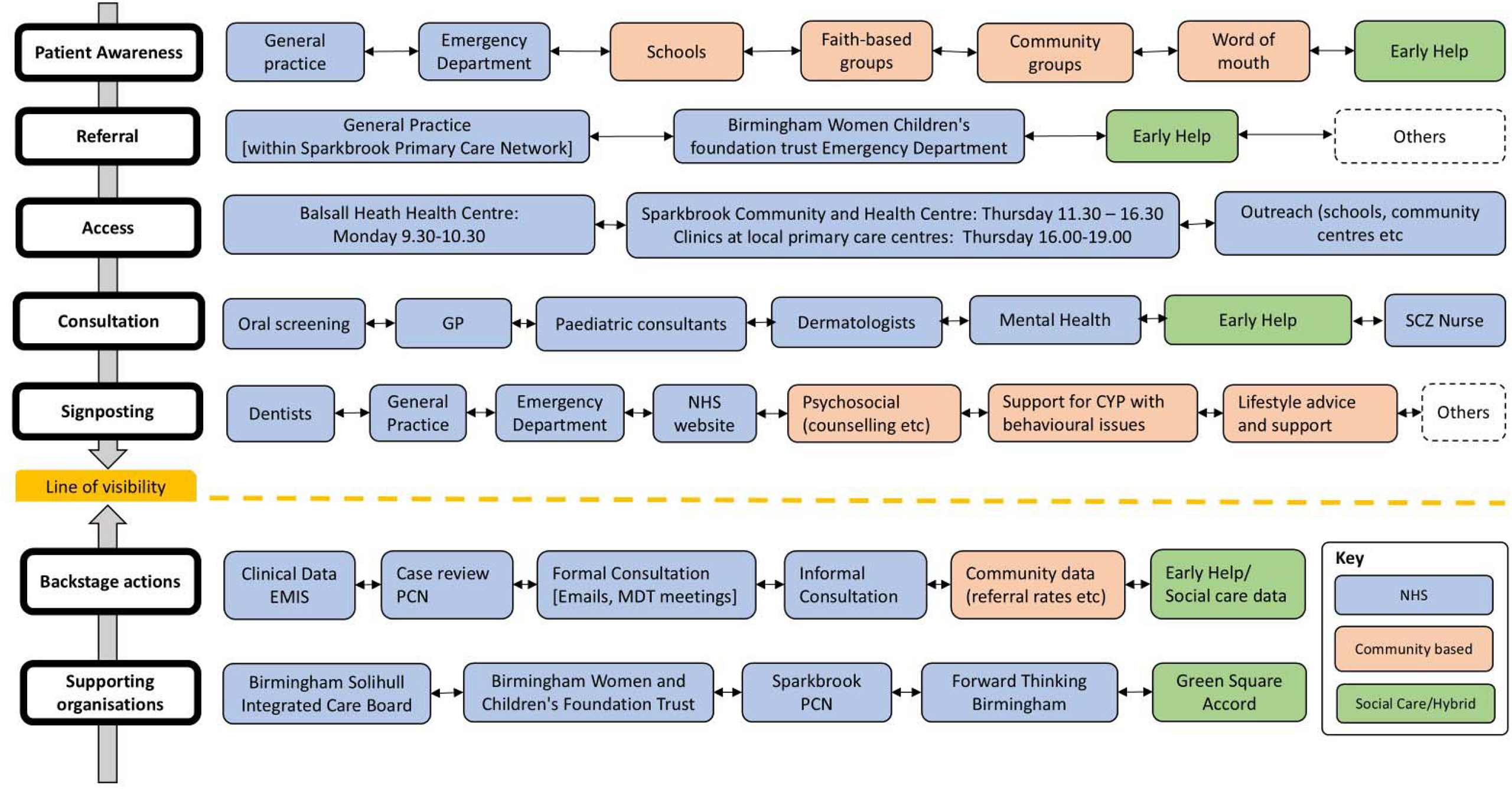
The Sparkbrook Children’s Zone service blueprint [10, 11]

## Methods

### Study Design

The study used a series of focus groups conducted with families that have used the SCZ. The data were analysed using a directed content approach to populate Sekhon’s theoretical framework of acceptability (TFA) [13, 14]. Ethical approval was granted by London–Harrow Research Ethics Committee 25/PR/0168.

### Setting/recruitment

The SCZ is based in the Sparkbrook & Balsall Heath East ward in Birmingham (UK). It is the second most populous ward in the city, with a superdiverse, young population, that has the second highest level of deprivation in the city, with high unemployment rates and high levels of universal needs around housing, food, clothing [15]. It is disproportionately affected by childhood obesity, child criminal and sexual exploitation and has one of the highest levels of infant mortality in England [15]. Patients (over the age of 14) and their parents or guardians were eligible for inclusion in the study. The SCZ liaison officer (CL) approached potential participants in-person. All were supplied with a participant information sheet and given the opportunity to ask questions about their participation and then provided informed consent before the focus group commenced.

### Data Collection

Focus groups were led by [1^st^ Author] trained and experienced in qualitative research, who was unknown to participants prior to the study. A topic guide used was informed by the existing literature and previous qualitative work conducted with staff [10, 11]. Questions included how they became aware of the service, what they understood of its structure, its accessibility, and which elements worked well or might be improved. (A summary of the topic guide can be found in Supplementary File 1). The focus groups were conducted at two community centres in the locale as the SCZ clinic. An interpreter was present at each focus group. They were digitally audio-recorded, and transcribed verbatim by an authorised transcription service and the data managed using nVivo.

### Data analysis

Analysis was informed by the theoretical framework of acceptability (TFA) developed by Sekhon et al to explore acceptability of health care interventions from the perspectives of providers and patients [13]. It consists of seven domains: affective attitude, burden, ethicality, intervention coherence, opportunity costs, perceived effectiveness, and self-efficacy (see Figure 2). Together they describe whether those receiving (or delivering) an intervention consider it to be appropriate based on anticipated or experiential cognitive and emotional responses [13]. Its structured approach allows for the isolation of the various components and attributes of an intervention and helps predict sustained engagement and effectiveness. It has been successfully used in a range of settings and circumstances including the treatment of HIV [16], breast cancer [17] diabetes [18] and mental health [19].

**Figure 2.**
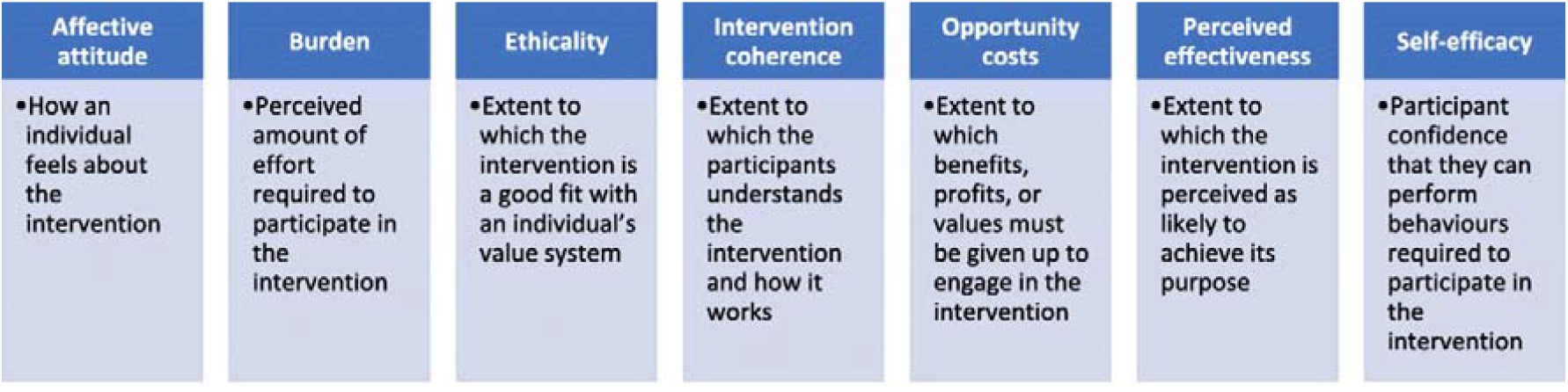
Summary of Sekhon’s Theoretical Framework of Acceptability

The first author [IL] independently coded each transcript fitting the data within each of the relevant domains of the TFA using a directed content analysis [14], following the unconstrained matrix approach suggested by Elo and Kyngäs to allow for the development and inclusion of emergent constructs or sub-constructs within the established framework [14, 20]. Ultimately the data was effectively allocated within four of the TFA’s domains: *Intervention coherence; Burden; Cultural Sensitivity*; and *Perceived effectiveness* with emergent constructs identified within each. The final allocation of data was consensually agreed by all authors.

## Results

### Participant characteristics

The intention was to run up to four focus groups but reasons for declining participation included a lack of ability to travel to the (community located) focus group venue, and caring and other familial responsibilities (the challenges of recruiting underserved groups for research studies are further explored in the *Strengths and limitations* section). Ultimately, a total of eight parents participated over two focus groups. The first consisting of 6 participants (plus one interpreter) lasted 47 minutes the second consisting of 2 participants (plus interpreter) lasted 24 minutes. There were five mothers and three fathers with a maximum of five children and a minimum of two. One participant did not disclose the total number of children. Because of the small number of participants and their potential identifiability we have minimised the description of each to gender and characteristics of their family (i.e. number of siblings) as further detail would risk compromising their confidentiality as per the ethical approvals.

**Table 1.** Participant characteristics.

| ***Participant ID*** | **Relationship to patient** | **Number of children** |
| --- | --- | --- |
| P01 | Father | Five |
| P02 | Mother | Three |
| P03 | Mother | Two |
| P04 | Mother | Four |
| P05 | Father | Two |
| P06 | Father | Undisclosed |
| P07 | Mother | Five |
| P08 | Mother | Two |

#### Qualitative findings

Below we present the qualitative findings in the four domains of the TFA describing the emergent constructs within each. These are summarised in Table 2 and presented below with exemplar quotes identified by their participant ID code and relationship to parent.

**Table 1.**
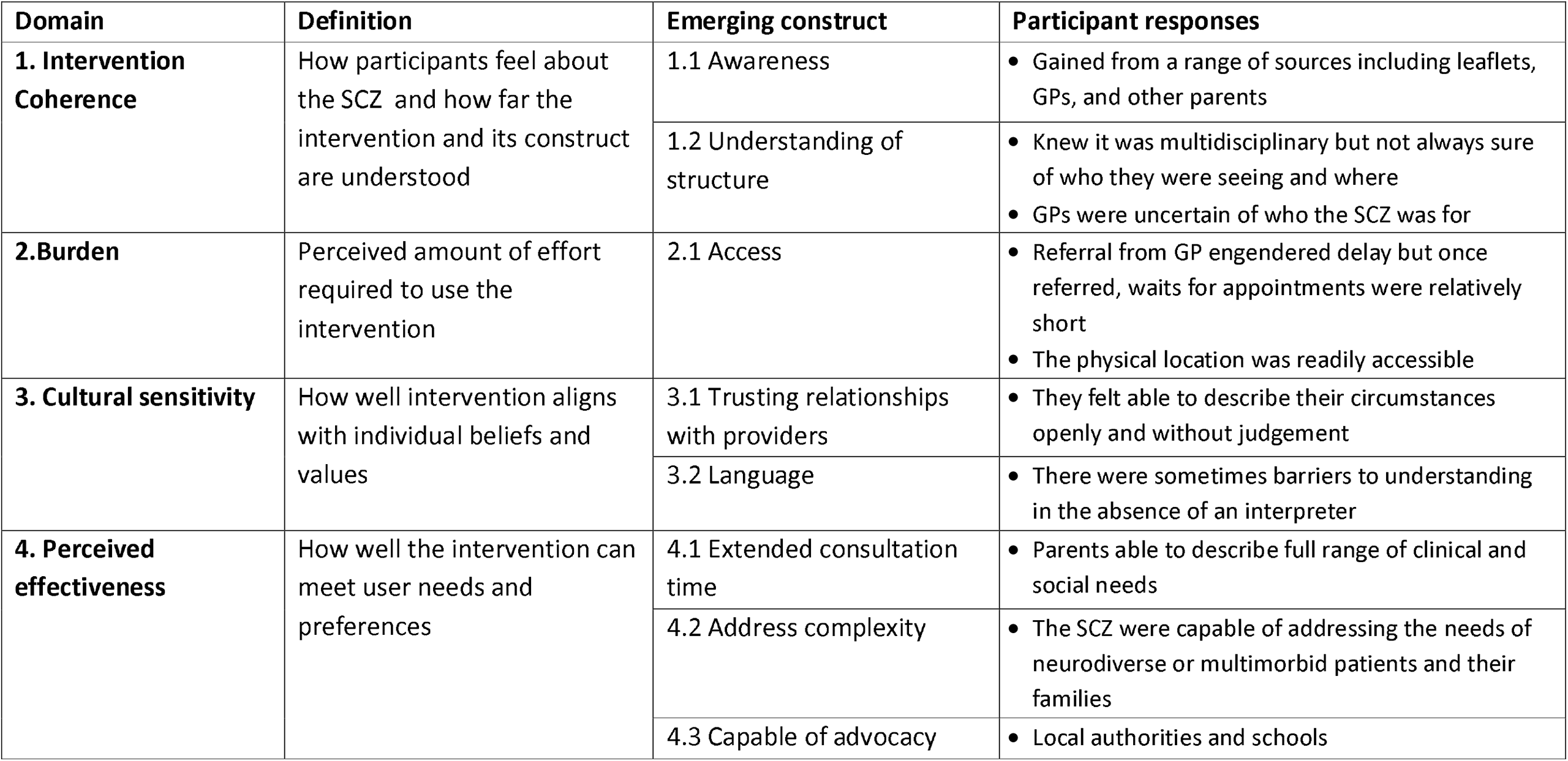
Summary of findings.

| **Domain** | **Definition** | **Emerging construct** | **Participant responses** |
| --- | --- | --- | --- |
| **1. Intervention Coherence** | How participants feel about the SCZ and how far the intervention and its construct are understood | 1.1 Awareness | • Gained from a range of sources including leaflets, GPs, and other parents |
|  |  | 1.2 Understanding of structure | • Knew it was multidisciplinary but not always sure of who they were seeing and where<br>• GPs were uncertain of who the SCZ was for |
| **2. Burden** | Perceived amount of effort required to use the intervention | 2.1 Access | • Referral from GP engendered delay but once referred, waits for appointments were relatively short<br>• The physical location was readily accessible |
| **3. Cultural sensitivity** | How well intervention aligns with individual beliefs and values | 3.1 Trusting relationships with providers | • They felt able to describe their circumstances openly and without judgement |
|  |  | 3.2 Language | • There were sometimes barriers to understanding in the absence of an interpreter |
| **4. Perceived effectiveness** | How well the intervention can meet user needs and preferences | 4.1 Extended consultation time | • Parents able to describe full range of clinical and social needs |
|  |  | 4.2 Address complexity | • The SCZ were capable of addressing the needs of neurodiverse or multimorbid patients and their families |
|  |  | 4.3 Capable of advocacy | • Local authorities and schools |

### Intervention coherence

Intervention coherence describes how far the intervention and its construct are understood by service users [13]. Parents described this within three constructs: how they became aware of the SCZ, their understanding of its structure in relation to its multi-disciplinarity, and the processes involved in follow-up.

#### 1.1 Awareness

The broad understanding of what the SCZ consisted of was derived from a variety of sources including their GP, leaflets at a local “play and stay” group, word of mouth from other parents, local events at a range of faith centres including both churches and mosques, and the SCZ’s outreach nurse that worked with local schools. A full explanation of what the SCZ delivered was provided once attending the zone (“…when we were at Children Zone, the entire process was explained to us…” *P06, Father*). However, it appeared that not all GPs in were fully aware of what the SCZ could provide:

> “I did originally ask for the Children Zone, but I feel like my GP sees them as a downgraded version of getting help or something, because for him they asked, “Okay, so what’s the issue?” So, I told the issues that I feel like he’s got autism, he’s got this - but they refused for me to see the Children Zone, and they said, “You need to see the doctor for this.”” *P06, Father*

#### 1.2 Understanding of structure

Families recognised the value of the clinical expertise they received (“the [Pediatric consultant] we saw at the Children Zone was extremely knowledgeable” *P06, Father*), as well as the practical advice and action of the family support officer:

> “As soon as [family support officer]…straight away she asked me everything, and then she was, “Okay [P03], I’m going to send you this, this, this, this, this…”” *P03, Mother*

Despite the understanding and appreciation of the multidisciplinary nature of the SCZ it left some unsure of who exactly they had seen and which organisation they were from:

> “Yeah. He’d walked us into the other room, and I think she did introduce who she was. But with him I just kept wondering afterwards was he a doctor, or was he something else?” *P02, Mother*

#### 1.3 Follow-up by SCZ

Participants described their lack of contact following their attendance at the SCZ (“…I’ve had no contact with them after that…*P02*). With some requesting further clarity as to the next steps including the timing of any follow-up:

> “I felt like…especially when we’re struggling with kids - that we can’t understand…it would have been nice to say, “Look, six weeks’ time, or even in a four to five months’ time we’ll give you a follow-up appointment, just to see how you’re getting on and what you need to do, and what you’ve tried and what you haven’t tried.”…” *P03, Mother*

### 2. Burden

The domain of Burden describes the perceived amount of effort required to use the SCZ [13] and this was described by the families in terms of their access to the SCZ.

#### 2.1 Access

Some participants were directly referred by their GP (“…My GP referred me because of the school was causing me distress about my daughter’s attendance…” *P03, Mother*), but for those that heard about it elsewhere they found the extra step of being referred by their GP frustrating, preferring to arrange a consultation directly with the SCZ:

> “It was really difficult to get an appointment through the GP though because they said ‘phone reception and ask for the Children’s Zone’, but they said, ‘No, you have to see the GP first, and then they will…[refer you]’…” *P02, Mother*

Once referred families described what they felt was a relatively short wait of one to two weeks for their appointment at the SCZ (“Yeah, so [once] I was referred to them. Got my appointment really quick, it wasn’t a long wait” *P03, Mother*). They were also satisfied that the location of the clinics was readily accessible on foot or by car (“…if you have car it’s easier, but if the worse the bus, or you can walk.” *P07, Mother*).

### 3. Cultural sensitivity

The domain of cultural sensitivity describes how well the intervention aligns with the beliefs and values of the individuals using it [13]. This was described in terms of the trusting relationships developed between SCZ providers and patients and their families and the challenges of native language.

#### 3.1 Trusting relationships

Families repeatedly described how they felt understood by SCZ staff, that the connection they made was personal. As this exchange within the first focus group exemplified:

> *P03 (Mother):* I think it’s more that they build a rapport.
>
> *P02 (Mother):* They don’t see you like a number, it’s really personalised.
>
> *P01 (Father):* The thing is they don’t just do the job, you know what I mean? They listen you, they care about you, you know? They give you the time, treat you like you’re a person they try to understand your problem…

This perception was supported by the willingness of SCZ staff to not only address the issues facing their child but also the broader challenges they faced as a parent:

> “Sometimes, I was so overwhelmed so just talking to her, she was not in rush at all. She used to listen me, sometime half an hour, 45 minutes, she was very patient, so helpful, you know what I mean?” *P01, Father*

#### 3.2 Language

The SCZ provides interpreters as part of its service, but they are not always available nor were they necessarily capable of speaking every language and dialect of what is a super-diverse patient population [15]. One participant explained through her interpreter, that she would take a friend or relative with her to consultations without which her understanding was patchy:

> “So [*P08’s]* daughter has actually got Down’s Syndrome, so her doctor referred her to Sparkbrook…and *[P08]* is just saying that she doesn’t really get to understand a lot, because she doesn’t understand English much… if someone is able to go with her to translate it there then she will understand better, but otherwise if she has to go alone then it’s just whatever she can make of it.” *P08, Mother via her interpreter*

### 4. Perceived effectiveness

The domain of perceived effectiveness refers to how well the intervention can meet user needs and preferences [13]. Specific to the SCZ three key elements emerged: the extended consultation time; its’ ability to address complexity of the patients and family’s needs; and the delivery of advocacy for patients.

#### 4.1 Extended consultation time

Participants repeatedly described the extended consultation allowed time to talk through a number of issues and challenges in contrast to the shorter ‘single problem’ consultation at their GP surgery:

> “…I’m happy because the nurse…I take long time, I ask a lot of things - but our GP? When you have appointment we must only *one* [symptom] talking about…I feel it’s good to get more help, more information.” *P07, Mother*

Although patients felt able to discuss a variety of inter-related clinical and social challenges, there was a structure to the provider response and the next steps:

> “…you’re just spilling your guts out, they allowed you to do it all, and then…they picked on certain topics. So, it was like, “Okay, say everything that you need to say, and let’s take it step by step.” And then it just went into bullet points, “Okay, you mentioned this, how should we cover that? What do you expect and what support you need?” *P03, Mother*

#### 4.2 Address complexity

Parents described how staff at the SCZ were willing to address more complex conditions and contexts than the usual care provided during a GP consultation. For example, a parent with an autistic child described how the SCZ provided signposting to further information and strategies that might be used with an autistic child, an element missing from consultations with their usual GP:

> “…when I met the Children Zone they were… it’s like so much information, and they were giving you in regarding to food habits, toiletry, even brushing their teeth - because autistic kids completely different understanding - linking you to websites and stuff, online courses, things like that. So, there’s a lot to it compared to when you go to the doctor and say to you… “Oh yeah, you’ve just got to monitor your child.”” *P03, Mother*

Another parent told through their interpreter how they had toured the NHS trying to find the right help for a disabled daughter who was suffering with depression until the SCZ provided the medical and social support they needed:

> “Because he met a lot of professionals, and they didn’t help, the help that he needs to have about his daughter, because she is disabled, and they have low income, and also the daughter she has depression… But when he met [family support officer] he asked first thing, “Please don’t leave me in the middle of the situation, so help me.” So she helped a lot, and also about the benefits, about the… that she needs physiotherapy…” *P05, Father* (via interpreter)

#### 4.3 Capable of advocacy

The ability of the family support officer to complement the clinical expertise of the SCZ was appreciated, particularly in terms of their advocacy with the local authority for housing:

> “… because she’s more the family support officer… it’s not just based about the kids, it’s also based about your own welfare…she knows I was on a council [home] waiting list, and she’s having a meeting with them, even though she signed me up [for a home] she’s, “I’m going to your housing officer…just to have a look that they’ve covered every base in your application form.” - and that’s something nobody ever does. Even if you go to Citizens Advice you won’t get that…, you phone allocations team you don’t get that, they’ll just say, “Yeah six months somebody will get back to you.” But she’s actually gone out of her way and said, “I’ll cover all these for you, you just concentrate on your kids…” *P03, Mother*

On another occasion the family support officer advocated for a family whose child was struggling at school, helping them better understand the circumstances of the child:

> “[family support officer] …she’s so good…she helped me with this while the school not understanding when I have explaining them that he’s going through that issue. But through Children Zone she went, she explained them, then she explained me, “That’s what I’m doing, don’t worry.” *P01, Father*

## Discussion

### Summary of findings

The patient voice is central to developing NMDTS that are appropriate and effective and this work has used the TFA to provide a structured understanding of the acceptability of a pilot NMDT in Birmingham, UK. In summary, by domain we found that in considering *Intervention coherence*, that individuals became aware of the SCZ through a range of sources, understanding that it was multidisciplinary if sometimes unsure of precisely the organisations involved; In *Burden* parents described the benefits of local collocated services and relatively short waits for an appointment once referred; In considering *Cultural sensitivity* parents described how they felt listened to by the care provider; finally in *Perceived effectiveness* they described the benefits of the extended consultation time, the SCZ’s ability to deal with complex patients and social contexts, and the benefit of the advocacy provided

### Specific findings

#### Awareness

The SCZ used a number of strategies to raise awareness including events at faith organisations, community locations, and local schools all previously successful in encouraging engagement in underserved communities [21]. The multi-disciplinarity of the service was also well understood and welcomed though there was some confusion over the exact speciality of those they were seeing and by their nature MDTs can reduce continuity of care and fragment relational continuity [22]. This has led to patients being treated by MDTs lacking awareness of the role of their (health) care providers or the identity of their main doctor [23]. It is important that patients remain aware of the structure and membership of their MDT in primary care settings with evidence that patients’ service awareness can influence engagement and outcomes [22]. The complexity of multi-agency MDTs can also cause uncertainty in the surrounding health care system, and one parent described how their GP failed to understand the content of the SCZ. If in the UK the concerted shift to NMDTs is to be successful then, as with any other fundamental shift in care delivery, it’s important that there is tsupport and engagement from the surrounding system [24].

#### Burden

There are a number of recognised barriers to accessing primary care in underserved populations including financial and temporal constraints and physical location [25]. Both the SCZ’s collocation and community setting facilitated access for parents echoing previous examples of where collocation in local communities improves access and reduces no-shows in underserved populations [26].

Parents felt that the wait of a week or two for an appointment was acceptable, though there were concerns over the delay in seeking a referral from their GP in the context of lengthening wait times for GP consultations [27]. Such waits can deter access amongst the underserved and there may be a role for direct online booking systems and symptom checkers to support access to NMDTs with the proviso that digital connectivity and literacy is appropriately supported [28].

#### Cultural sensitivity

Parents described the development of trusting, personalised relationships with their care provider. The consultation style of the SCZ follows many of the principles of the “inner consultation model”: making a connection; summarising key points; and ensuring parents were aware of next steps [29]. The personalised nature of the parent-provider relationship was also fostered by other recognised strategies including a caring communication style, and exhibiting an interest in the patient [30]. Building trust is of particular value in underserved populations that often have poor experiences of mainstream healthcare. In the UK the Royal College of General Practice has described the value of this relationship-based care, establishing rapport and empathy with patients [31] and there is growing use of similar non-transactional approaches that prioritises trust and ongoing care in the United States [32].

Despite the ability of SCZ staff to foster trusting relationships with families, familiar issues around interpretation were described. Its known that language barriers can compromise quality of care, patient safety and health outcomes [33]. The SCZ did provide interpreters and employed multi-lingual members of staff but the local population speaks some 70 languages presenting logistical issues witnessed across primary care [34].

#### Perceived effectiveness

Previous work has reported how additional time is required in complex consultations with underserved populations [35]. Parents described how the extended consultation time allowed for discussion of their family’s complex medical and social needs. There is already evidence that longer consultations are associated with better health outcomes [36] (in contrast to shorter contact times that can lead to more frequent visits [37]). As parents were able to discuss an unlimited number of issues, this provided a more holistic image of the patient than allowed by existing recommendations in the UK for one problem per appointment.

Many healthcare systems are configured such that General Practitioners (GPs) have an important role in identifying autism in children and managing their care [38]. Despite this GP knowledge and experience in identifying autism and managing care for children with the condition is varied [39], with many employing the gatekeeping principles of ‘wait and see’ [38]. This chimes with the experience of parents of neurodiverse children in Sparkbrook prior to attending the SCZ and previous reports that the needs of neurodiverse children are dismissed, minimised, or otherwise unacknowledged by frontline healthcare professionals [40].

Parents were grateful for the advocacy provided by the family support officer and in the UK, where there are some £23 billion in unclaimed benefits [41], there is a growing recognition of the importance of advocates in health and social care in the UK, including recommendations by the National Institute for Clinical Excellence [42]. In our qualitative study with SCZ staff, we also noted that families found accessing social support was less stigmatising when delivered in a children’s clinic {Litchfield, 2025 #50}. The ability of advocacy to support patients’ voices and mobilise resources, including challenging decisions being made by councils, was observed in the SCZ and elsewhere [42]. However, more work is needed to understand how advocates can most effectively be included within primary care teams [43].

### Strengths and limitations

The emerging constructs highlighted key advantages and potential challenges associated with NMDTs for children, particularly those from underserved populations. Populating the TFA by using an open matrix approach with two researchers coding independently bolstered analytical credibility and trustworthiness [14]. The number of participants were lower than anticipated and although this may be considered to limit broader generalizability, consensus theory tells us that a smaller number of ‘experts’ with shared knowledge of a tightly defined topic are sufficient to describe common experiences and values [44]. However, the data are able to offer valuable insight transferable to similar populations and contexts without the collection of superfluous data [45].

## Conclusions

NMDTs are being rapidly introduced into health and social care in the UK despite a lack of corresponding evidence. This work has offered valuable insight into parent perspectives of an NMDT for CYP in a diverse and economically disadvantaged ward in Birmingham. Parents appreciated the collocated and community-sited premises, the provision of relationship-based care, advocacy and support for the clinical needs of their child and broader social circumstances. However, more work is needed to ensure that the structure of the MDT is thoroughly understood, access is streamlined and allowance is made for linguistic diversity.

## Data Availability

All data included in the study are available upon reasonable request and within the stipulations of the ethical approval granted

## Acronyms

CYP: Children and Young People
NMDT: Neighbourhood Multidisciplinary Team
NHS: National Health Service
SCZ: Sparkbrook Children’s Zone
TFA: Theoretical Framework of Acceptability
UK: United Kingdom
VCFSE: Voluntary Community, Faith and Social Enterprise

## Authors statement

### CRediT authorship contribution statement

Ian Litchfield: Conceptualization, Methodology, Investigation, Data curation, Formal analysis, Writing-original draft, Writing – review & editing

Fran Dutton: Validation, Writing – review & editing

Lorraine Harper: Conceptualization, Funding acquisition; Writing – review & editing

Simarjeet Kaur: Project Administration, Validation, Writing – review & editing

Chiara Luxmoore: Project Administration, Validation, Writing – review & editing

Lubna Rahman: Project Administration, Writing – review & editing

Caroline Wolhuter: Validation, Writing – review & editing

Chris Bird: Conceptualization, Funding acquisition; Validation, Writing – review & editing

### Ethical approval

Ethical approval was granted by the University of Birmingham’s Science, Technology, Engineering and Mathematics (STEM) ethics committee (ERN_22-1839)

### Availability of data and materials

The datasets generated and/or analysed during the current study are not publicly available as the consent did not include the dissemination of the data for use beyond this study.

## Supplementary File 1 Summary interview topic guide

*Introductory question*

- Can you all please tell me a bit about yourselves (for example, how long have you lived in your neighbourhood? How many children do you have?)

*Thinking about your awareness of the service…*

- How did you hear about the service?
- What did you understand about what it offered?
- Was the reason for your referral explained?

*Thinking about your consultation…*

- How easy was it to get to the premises and find the service?
- Who did you see?
- Were you signposted to other care providers within the building?

*Thinking of next steps…*

- Following your appointment, did you attend or were you directed to any other services

*Thinking of your overall experience…*

- How did you find the service provided by SCZ?
- What aspect went well or might be changed to make it easier using the service?

